# Dynamic Changes in Distribution of Hydrocodone and Oxycodone in Florida

**DOI:** 10.1101/2022.08.22.22279071

**Authors:** Elena L. Stains, Akshay C. Patel, Joseph D. Hagedorn, Jay P. Solgama, Kenneth L. McCall, Brian J. Piper

**Author notes:** Akshay C Patel, Department of Medical Education, Geisinger Commonwealth School of Medicine, 525 Pine Street, Scranton, PA 18509, United States, or.

## Abstract

**Purpose:** The opioid epidemic in the United States began with medical providers over-prescribing opioids. Florida, which led the country in opioid-prescribing physicians, was unique during this period because of its lax prescribing laws and high number of unregulated pain clinics. Here we address the difference in distribution rates of oxycodone and hydrocodone across Florida counties during the peak years of the opioid epidemic.

**Methods:** Washington-Post and the United States Drug Enforcement Administration’s Automation of Reports and Consolidated Orders System (ARCOS) databases provided longitudinal oxycodone and hydrocodone prescription data in grams per county (2006-2014) and statewide (2006-2021). Grams of oxycodone and hydrocodone were converted to morphine milligram equivalents (MME) for comparison.

**Results:** There was a steep increase in oxycodone from 2006 to 2010, with subsequent decline. Hydrocodone distribution decreased slightly from 2006 to 2014. In peak year, 2010, the average MME per person across all counties in Florida was 729.4, a 120.6% increase from 2006. The three individual counties with the highest MME per person in 2010 were Hillsborough (2,271.3), Hernando (1,915.3), and Broward (1,726.9) and were significantly (p < .05) elevated relative to the average county. MME per person was highly correlated (r=0.91) with MME per pharmacy, therefore in most counties, both values rose together.

**Conclusion:** The novel data demonstrated pronounced differences in opioid distribution, particularly oxycodone, between Florida counties during the height of the opioid epidemic. Legislative action taken between 2009 and 2011 aligns with the considerable decline in opioid distribution after 2010.

**Key Points:**

- The 2000s saw a rise in opioid use, misuse, and overdose deaths across the United States, especially in Florida.
- Morphine Milligram Equivalents (MME) of oxycodone increased 230.2% in Florida from 2006 to the peak distribution year, 2010.
- Average MME per person in the state increased 120.6% from 2006 to 2010, while some counties’ MME per person rose over 150%.
- Eleven counties’ average MME per person were significantly higher than the state’s average.
- There was considerable variation between counties—16.6x higher MME per person in Hillsborough than in Liberty in 2010.

**Plain Language Summary:** The opioid epidemic in the United States began with medical providers over-prescribing opioids. Florida, which led the country in opioid-prescribing physicians, was unique during this period because of its lax prescribing laws and high number of unregulated pain clinics. Here we address the difference in the distribution of two popular opioids, oxycodone, and hydrocodone, across Florida counties during the peak years of the opioid epidemic. The United States Drug Enforcement Administration’s Automation of Reports and Consolidated Orders System (ARCOS) database obtained by the Washington Post provided oxycodone and hydrocodone data from 2006 to 2014. Grams of oxycodone and hydrocodone were converted to morphine milligram equivalents (MME), a standardized opioid measurement, for comparison. There was a steep increase in oxycodone from 2006 to 2010, followed by a decline. Hydrocodone decreased slightly from 2006 to 2014. In the peak year, 2010, the average MME per person across all counties in Florida was 729.4, a 120.6% increase from 2006. The three counties with the highest MME per person in 2010 were Hillsborough, Hernando, and Broward and were significantly (p < .05) elevated relative to the states average. The data demonstrated major differences in opioid distribution, particularly oxycodone, between Florida counties during this period.

## INTRODUCTION

Opioid distribution rapidly accelerated in the United States (US) in the 2000s. In 2013 and 2014, there were 10.7 million people who reported abusing pain relievers in the past year, with one-quarter (25%) of the pills being sourced directly from physicians^1^. Oxycodone and hydrocodone product misuse was particularly common among pain reliever abuse^2^. Deaths due to opioid overdose increased from over 8,000 in the year 2000 to over 46,000 in 2018^3^. In fact, the burgeoning trend of medical providers over-prescribing of opioids to patients during this period is often cited as a driver of the Opioid Crisis in the United States. Opioids are commonly prescribed to control moderate to severe acute pain^4^. Physicians usually prescribe these narcotics after major surgery or severe injury, or even for chronic pain^4,5^. Some patients develop a tolerance for the medication, requiring a higher dose to achieve the same level of pain relief. Patients also face the risk of forming physical dependence which may be accompanied by withdrawal symptoms if they lose access to the medication.

Oxycodone and hydrocodone were discovered in 1920 and 1916 respectively in Germany^6^. The mechanisms of action of these two drugs are similar. Both drugs are agonists of mu, kappa, and delta opioid receptors. As these receptors are G-coupled proteins, the interaction results in the exchange of GDP for GTP, leading to a decrease in intracellular cAMP. This decrease results in the inhibition of nociceptive neurotransmitters such as acetylcholine, dopamine, GABA, and substance P. Both hydrocodone and oxycodone are administered orally. Both opioids are often prescribed as combination medications with either acetaminophen, aspirin, or ibuprofen. Long term use of hydrocodone may cause QT wave elongation, constipation, and hypotension. Oxycodone use, on the other hand, commonly results in respiratory depression, pruritis, and nausea. Both of these Schedule II opioids come with labeled warnings for the risk of addiction, and respiratory depression, as well as a warning stating the medications should not be taken while pregnant. On average, the price of a 10 mg capsule of hydrocodone is about $8.64, versus oxycodone which cost about $0.58 for a 5 mg capsule (Supplemental Table 1).

One of the key states leading the US opioid epidemic was Florida. The preponderance, 98 of the United States’ top 100 opioid-prescribing physicians in 2010, operated out of Florida^7^. During this year, the state reached a peak in opioid distribution and ranked 13 out of 50 states in opioid overdose death rate^8,9^. To deal with the erupting opioid crisis, the Florida government began to address the high distribution of opioids in the state by creating the Prescription Drug Monitoring Program in 2011. Two years later, Florida signed into law HB7095, which aimed to control rogue pain management clinics in the state by limiting a provider’s ability to dispense opioids directly^10^.

Here we explore opioid distribution across Florida counties during the peak distribution years of the opioid epidemic. Although there has been some prior research on this topic,^8^ this is the first examination of this state using a novel database which was obtained by the Washington-Post that provides county-level information^11,12^.

## METHODS

### Procedures

Data obtained from the United States Drug Enforcement Administration’s Automated Reports and Consolidated Orders System (ARCOS) and by the Washington-Post provided longitudinal oxycodone and hydrocodone distribution^12^. Data obtained by the Washington Post was converted to grams of opioids per county for 2006 to 2014. However, we also include data from the standard ARCOS^13^ for 2006 to 2021, to provide further insights into hydrocodone and oxycodone distribution beyond 2014. The ARCOS data reports opioids distributed to several business types (provider, pharmacy, hospital, etc.), while the Washington Post data includes only opioids distributed to pharmacies.

Grams of oxycodone and hydrocodone were each converted to morphine milligram equivalents (MME) and totaled^14^. For comparison, the daily dose of oxycodone or hydrocodone for moderate pain after laparoscopic surgery is 15 to 20 mg. ARCOS has been validated by comparing it to a state prescription drug monitoring program and the result was r = 0.985^15^. The Washington-Post ARCOS database has been used in previous research^16–18^. There was a high correlation between the traditional ARCOS and the Washington-Post ARCOS (Supplemental Figure 1). Each county’s (Supplemental Figure 3) total MME was then divided by county population during the given year according to the American Community Survey to obtain MME per person^19^. MME per person across counties and study years was plotted to assess peak distribution year. We then designated three counties as the “top three counties” and three counties as the “bottom three counties” based upon their MME per person during the peak year, 2010. We created heat maps to visually represent each county’s MME per person and MME per pharmacy using RStudio. Procedures were approved by the IRBs of the University of New England and Geisinger.

### Data-analysis

A 95% confidence interval (1.96*SD) was determined and counties were considered statistically different if they fell outside of this range. This was used to evaluate each county’s average MME per person across study years as well as the MME per person in the peak year.

## RESULTS

### ARCOS 2006-2021

ARCOS data shows a sharp increase (+215.3%) in oxycodone distribution from 2006 to 2010. There were 18.72 metric tons of oxycodone distributed in the state in 2010. Oxycodone distribution decreased (−69.1%) from 2010 to 2014, and remained low, despite a small rebound from 2016 to 2017. Hydrocodone decreased 58.5% from 2006 to 2021 (Figure 1A).

**Figure 1.**
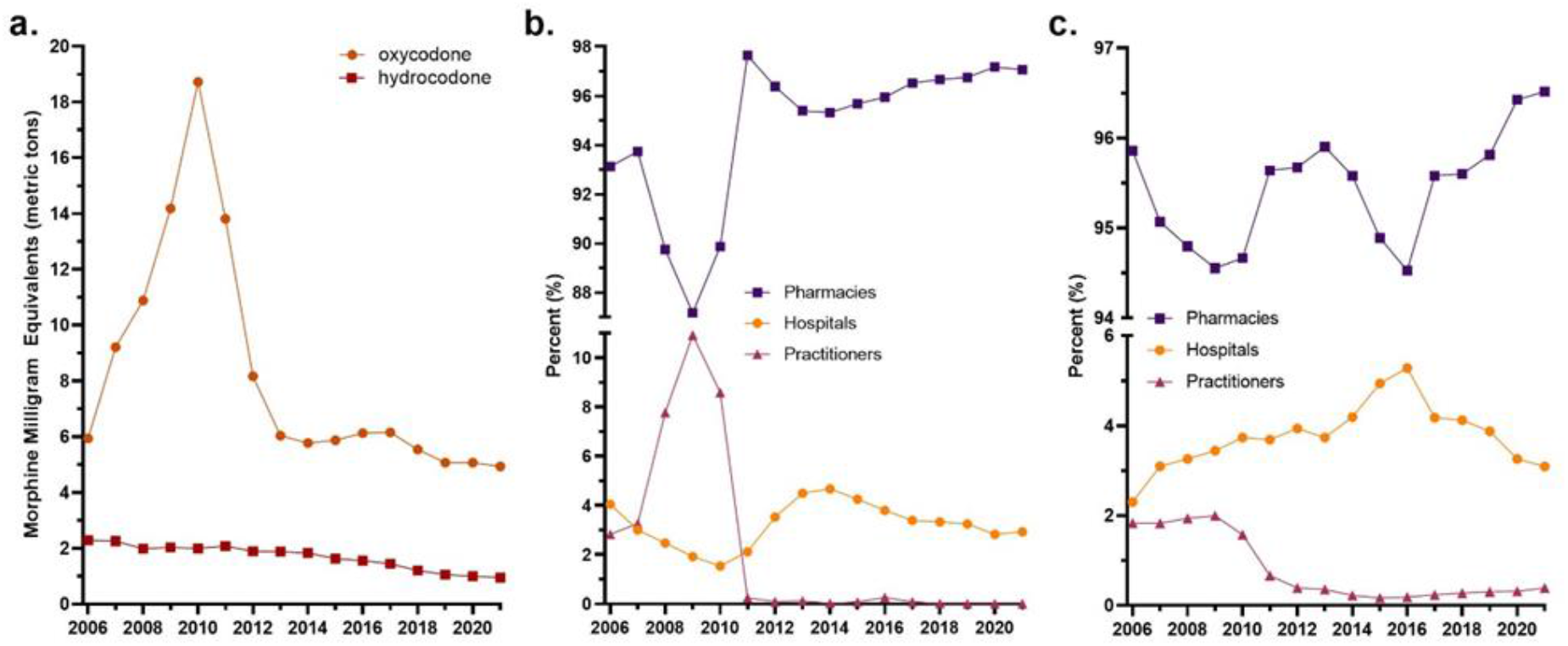
Drug Enforcement Administration’s Automated Reports and Consolidated Orders System weight of oxycodone and hydrocodone distributed in Florida from 2006 to 2021 (a). Percent of oxycodone (b) and hydrocodone (c) distribution by business activity.

Of the total weight of oxycodone distributed in 2006, 2.83% was distributed by medical practitioners. This percentage increased to 10.9% in 2009, and then dropped to nearly zero by 2011 (Figure 1B). As for hydrocodone, 1.84% was distributed by practitioners in 2006. This number also decreased after 2009, remaining under 0.4% through 2021 (Figure 1C).

### Washington Post ARCOS 2006-2014

The state distributed a total of 103,315.43 MMEs in kilograms of hydrocodone and oxycodone from 2006 to 2014. Florida saw a steady increase in MMEs of oxycodone (+230.2%) from 2006 until it peaked in 2010. However, the MMEs of hydrocodone decreased (−11.5%) from 2006 to 2010. A total of 19,958.85 MMEs were distributed in 2010. Distribution subsequently declined for oxycodone (−70.2%) and hydrocodone (−7.2%) from 2010 to 2014.

When all counties were averaged, there was more than a three-fold rise in oxycodone from 2006 to 2010 (Figure 2A). While the total amount of oxycodone in Florida increased from 2006 to 2010, there was great variability among counties. Dixie County distributed only 2.95 MME kg in 2006, and decreased distribution -10.2% to 2.65 MMEs by 2010. However, most counties increased their distribution substantially. Broward County’s distribution rose 147.5% from 1,222.78 MMEs in 2006 to 3,026.98 MMEs in 2010.

**Figure 2.**
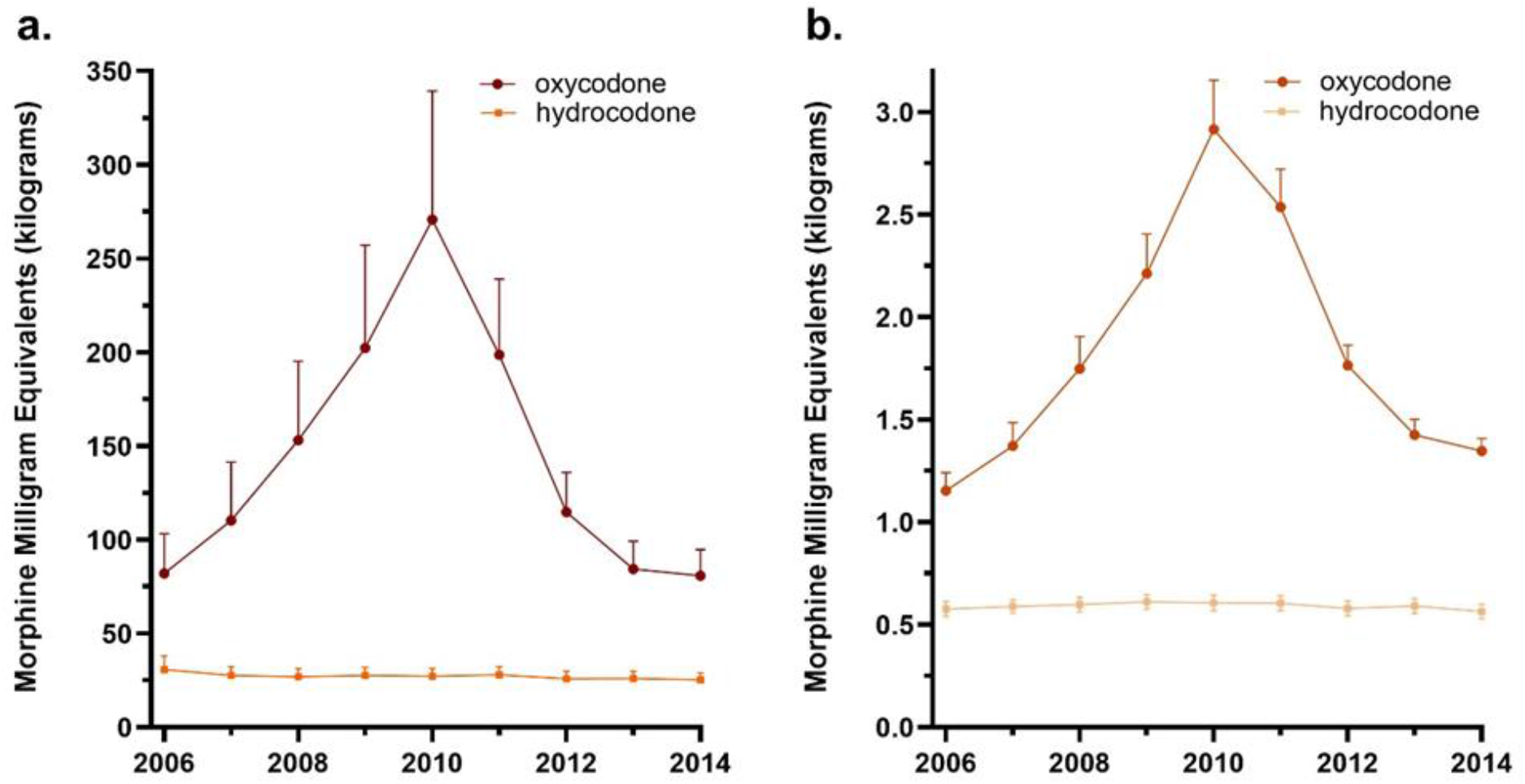
Morphine milligram equivalent (MME) per capita (a) and MME per pharmacy (b) across all Florida counties for oxycodone and hydrocodone during 2006 to 2014 as reported to the Washington Post/Drug Enforcement Administration’s Automated Reporting and Consolidated Orders System (ARCOS).

The MMEs per pharmacy followed a similar, yet less pronounced, pattern. From 2006 to 2010, there was a 152.7% increase in MMEs of oxycodone, and a 53.8% decrease after 2010. Hydrocodone declined slightly (−2.1%) from 2006 to 2014 (Figure 2B).

Florida’s mean MME per person across study years was 490.0. Eleven counties’ mean MME per person were significantly higher than the state’s (490.0): Baker (750.1), Broward (891.9), Charlotte (863.6), Hernando (1019.2), Hillsborough (1110.0), Okeechobee (844.2), Palm Beach (786.4), Pasco (940.5), Pinellas (915.8), Putnam (812.3), and Sarasota (769.5).

In 2010, the average MME per person in milligrams across the state was 729.4. This was a 120.6% increase from 2006, when the average MME per person in Florida was 330.7. The three individual counties with the highest MME per person in the peak year were Hillsborough (2,271.3), Hernando (1,915.3), and Broward (1,726.9) which were significantly (p < .05) elevated relative to the state’s average in 2010. The three counties with the lowest MME per person in peak year were Dixie (162.0), Lafayette (154.8), and Liberty (136.8). However, none of the three were significantly different from the state average in 2010 (Figure 3A).

**Figure 3.**
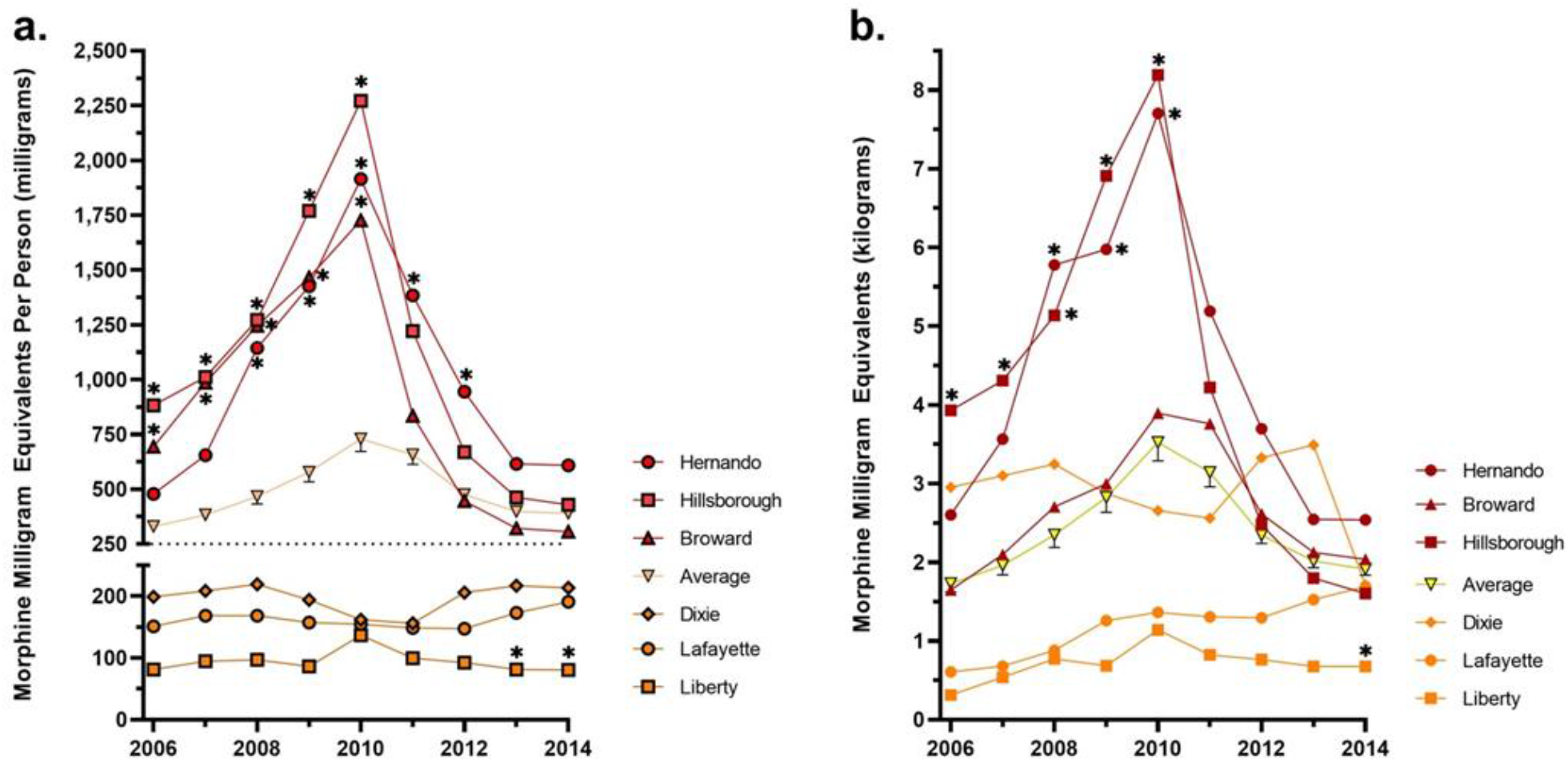
The morphine mg equivalent (MME) per person (a) and per pharmacy (b) in the top three (Hernando, Hillsborough, Broward) and bottom three (Dixie, Lafayette, Liberty) counties relative to Florida’s average as reported to the Washington Post/Drug Enforcement Administration’s Automated Reporting and Consolidated Orders System (ARCOS). Those counties that were statistically different (p < .05) from the mean are marked with an asterisk.

MME per pharmacy again follows a similar pattern. When comparing our top three and bottom three counties, most display the same pattern, with the exception of Broward and Dixie (Figure 3B). The MME per person correlated highly (r = 0.91) with the MME per pharmacy (Figure 4).

**Figure 4.**
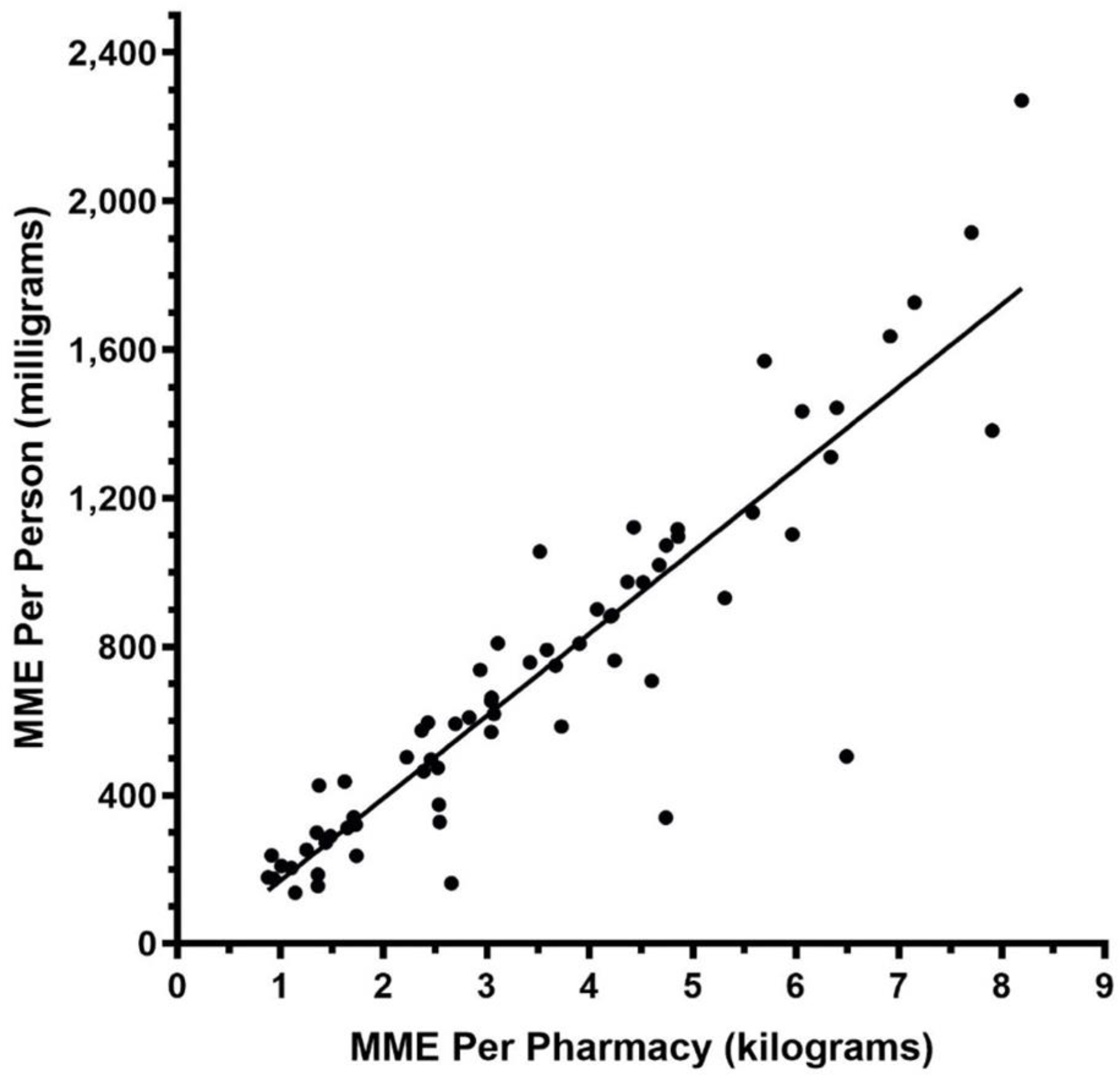
Correlation (r = .0.91) between MME per person and MME per pharmacy in the peak year, 2010 as reported to the Washington Post/Drug Enforcement Administration’s Automated Reporting and Consolidated Orders System (ARCOS).

**Figure 5.**
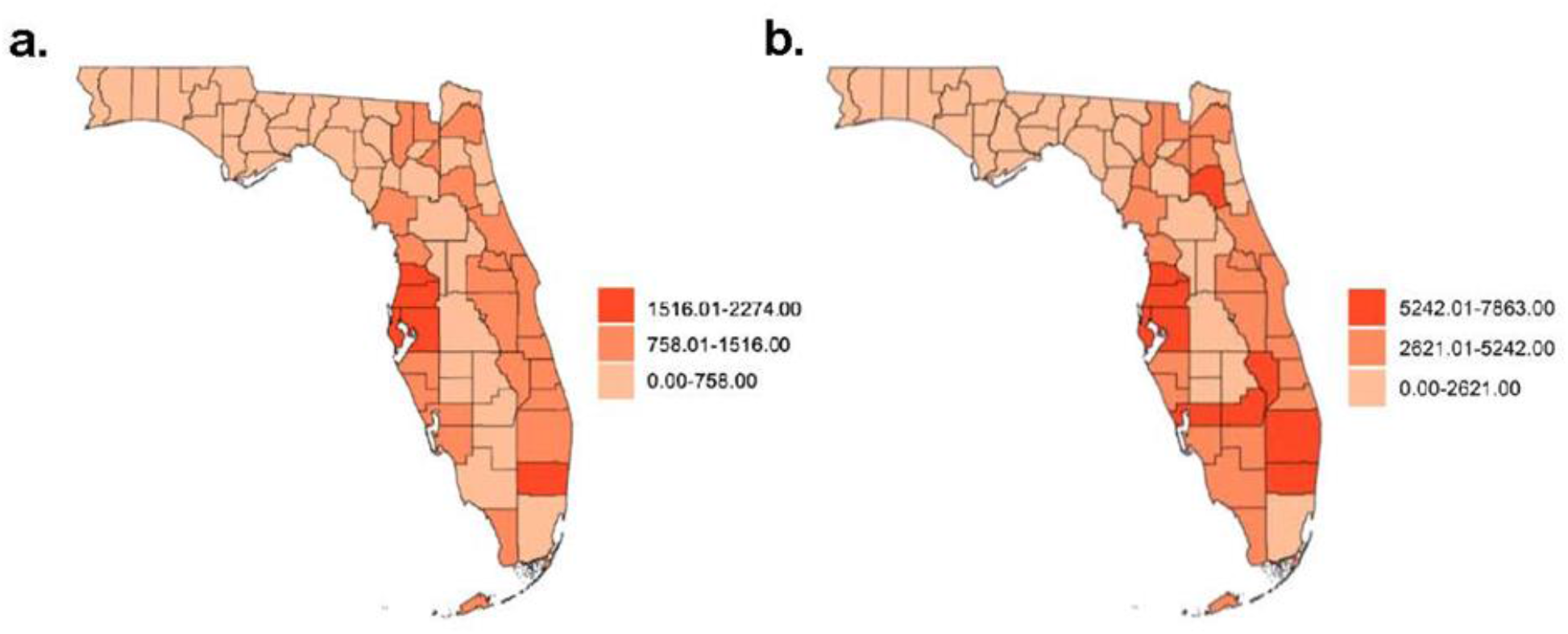
Heat maps representing the morphine mg equivalent (MME) per person (a) and MME per pharmacy (b) per county in 2010 as reported to the Washington Post/Drug Enforcement Administration’s Automated Reports and Consolidated Orders System.

## DISCUSSION

There are several key findings regarding oxycodone and hydrocodone distribution in Florida. There was a steep increase in distribution of these Schedule II opioids in the sunshine state from 2006 to 2010, followed by a similarly large drop-off. Raw MME distribution more than doubled. The pronounced increase in the total MME was attributable to a rise in oxycodone as hydrocodone distribution declined over this period.

The average MME per person across the state rose in a similar pattern to raw MME. However, when broken down by county, we saw that while some counties followed this uptick, others remained steady with very limited distribution. A few counties, namely Broward, Hillsborough, and Hernando, saw a peak in MME per person far above the state’s average. This pattern seems to have some regional patterns as Hillsborough, Hernando, and Pasco are adjacent to each other on Florida’s west coast in the vicinity of Tampa^20^.

The final noteworthy finding is the great decline in MME and MME per person in the state after the peak in 2010. The health policies of the state at the time may offer some insights into why these numbers dropped (Supplemental Figure 2). The sharp increase in opioid distribution in 2010 is thought to be a consequence of the many infamous “pill mill” physicians in Florida at that time^7^. In 2009, as opioid use was rising, the state passed legislation creating a prescription drug monitoring program (PDMP) known as E-FORCSE (Electronic-Florida Online Reporting of Controlled Substance Evaluation Program) to make it more difficult for physicians to maliciously prescribe controlled substances. The program, which was not implemented until 2011, required dispensers of controlled substances to file a report each time a Schedule II, III, IV, or V substance is given to a patient. It became the responsibility of the substance dispenser to check the history of the patient on the PDMP to further ensure abuse was not detected. To deal specifically with the vast number of pill mill physicians in Florida, the United States DEA launched a multi-agency operation called Operation Pill Nation to seize clinics, arrest physicians and clinic staff, and in some instances seize assets such as exotic cars and weapons^21^. Immediately after the raids is when Florida legislators passed HB7095, barring physicians from dispensing schedule II or III drugs directly out of their office. A key lesson from Florida’s experience was the importance of pharmacists in providing a crucial step in legitimate pain management.

More recently, Florida continued to decrease opioid related deaths through new health and safety bills. HB21, passed in July of 2018, resulted in the decrease of long-term opioid prescriptions, with a rise in 1 to 3 day prescriptions^11,22^. After witnessing the impacts of pill mill pain clinics, many other states implemented laws to control the prescription rates in their clinics. Other research has found modest or non-existent benefits of opioid prescribing laws^23,24^. The results from this research highlight the importance of legislative regulation on controlled substances such as hydrocodone and oxycodone, particularly eradicating “pill mills”. However, despite prior excesses in prescription opioid prescribing, Florida ranked only #40 out of 50 states in the US in 2019 for methadone treatment programs^25^.

The top three counties, Hillsborough, Hernando, and Broward, are home to the cities of Tampa, Spring Hill, and Fort Lauderdale, respectively. The bottom three counties, Dixie, Lafayette, and Liberty are located in the north of Florida and contain Cross City, Mayo, and Bristol, respectively. Hillsborough and Broward counties each have populations of over one million, while Dixie, Lafayette, and Liberty counties have some of the lowest populations in the state at under 18,000 people. The top three counties have generally higher median incomes ($60,566, $50,280, and $60,922) than the bottom three counites ($41,674, $51,734, $39,121). The top counties also have lower poverty rates (11.9%, 12.5%, 11.0%) than the bottom counites (23.2%, 20.7%, 21.2%)^26^.

The top three counties were likely distributing such disproportionate numbers of opioids because they were the sites of these “pill mill” clinics that were overhauled by the DEA. One infamous pain clinic arrest was that of Sylvia Hofstetter, who operated pill mills in Broward County and Tennessee, helping to distribute more than 11 million pills of oxycodone, oxymorphone, and morphine^27^. The Florida Department of Law Enforcement reported in 2010 that Tampa alone contained 103 pain clinics^28^. Similarly, Broward County had over 150 pain clinics in 2009^28^. Even some pharmacies were responsible for the high distribution of opioids—53 arrests were made at Glory Pharmacy in Hernando County, when police found they were not only accepting fraudulent scripts for controlled substances, but the owners were trafficking oxycodone pills by the hundreds^29,30^.

A key question is why the increase in oxycodone and not hydrocodone? The 2015 National Survey on Drug Use and Health found that hydrocodone products were the most commonly used and misused pain relievers in the United States, contrary to what Florida’s data indicates^2^. However, a factor that may have affected providers in Florida is the FDA’s 2013 recommendation to change hydrocodone products from Schedule III controlled substances to Schedule II^31^. This announcement and anticipation of hydrocodone rescheduling may have contributed to providers choosing other, less scrutinized pain relievers, like the previously uncontrolled, unscheduled tramadol^32^.

The standard ARCOS database which reports on Schedule II and III substances with limited geographic resolution has been used in many prior reports^8,15,23-25^. This report with the Washington Post ARCOS adds to a much more limited database of peer-reviewed investigations^33^.

### Limitations

This study focused on characterizing the opioid distribution pattern for Florida over a fifteen-year period (2006 – 2021) using two similar datasets, each with their own strengths. The traditional ARCOS is updated approximately every six months but does not provide for geographic resolution beyond that of the 3-digit zip code. The Washington Post ARCOS does not provide any data beyond 2014 but can be analyzed to the level of the individual county or pharmacy. The correlation between the two data sets was strong, but not perfect (r^2^= 0.98, Supplemental Figure 1). The Washington-Post ARCOS data was processed such that they removed repeated and null data sets before publishing^34^. As a part of the cleaning of the data, the Washington Post removed shipments of opioids that did not make it to the consumer^34^. The non-perfect association is likely due to these refinements in the Washington Post ARCOS.

This study examined only the pattern for oxycodone and hydrocodone, which are generally the most prescribed opioids for pain as prior investigations have evaluated other opioids and states^8,15,35^. Further investigation may reveal the possible socio-economic, geographic, and political impacts that caused certain counties to be greatly affected by the increased prescriptions compared to others.

In conclusion, this study used a novel database to evaluate the pronounced temporal and geographic variation in oxycodone prescribing in Florida. These findings may be valuable for preventing future iatrogenic opioid epidemics in the US or globally.

## Supporting information

Supplemental Table 1. Properties of hydrocodone and oxycodone.36

## Data Availability

All data produced in the present study are available upon reasonable request to the authors

