## Supplemental Table 1. Properties of hydrocodone and oxycodone.36 for "Dynamic Changes in Distribution of Hydrocodone and Oxycodone in Florida"

**SUPPLEMENTAL MATERIALS**

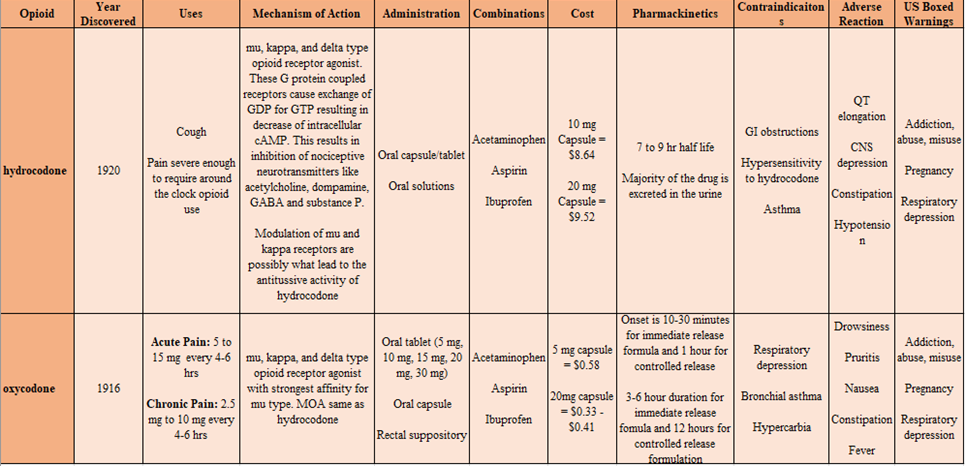

**Supplemental Table 1.** Properties of hydrocodone and oxycodone.^36^

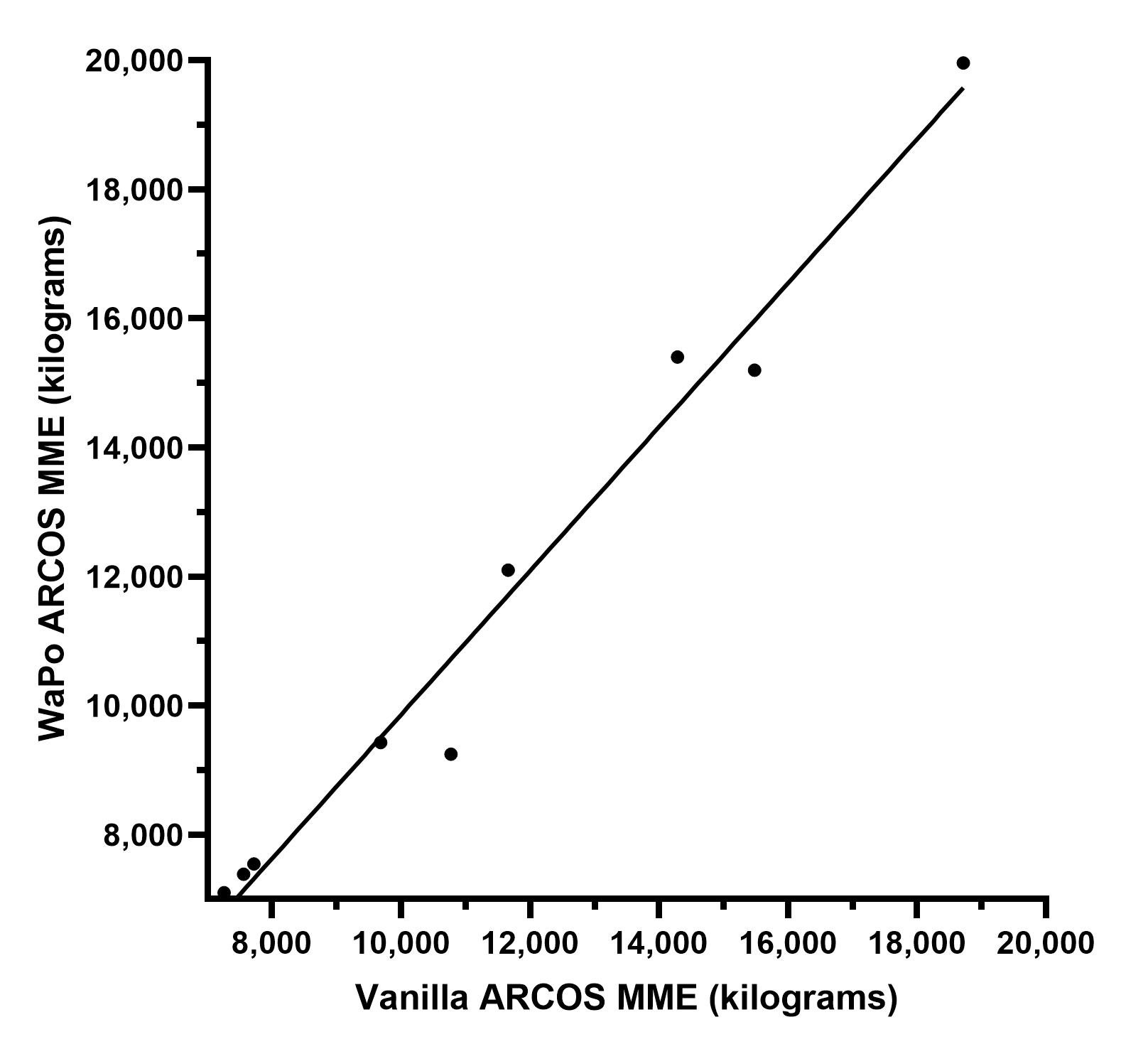

**Supplemental Figure 1.** High correlation (r = .99 p < .001) between the standard Automated Reports and Consolidated Orders (ARCOS database for pharmacy distribution of oxycodone and hydrocodone and the Washington Post (WaPo) ARCOS database. Each data point represents a single year from 2006-2014.

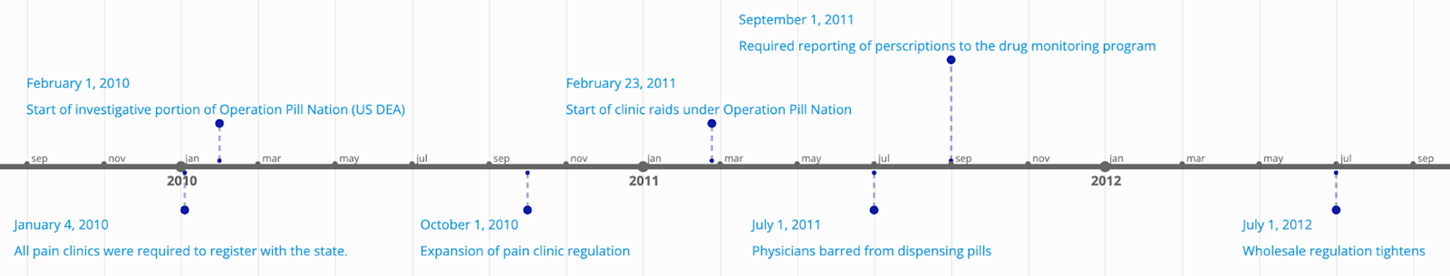
**Supplemental Figure 2**. Timeline of Florida laws from 2009 to 2012.

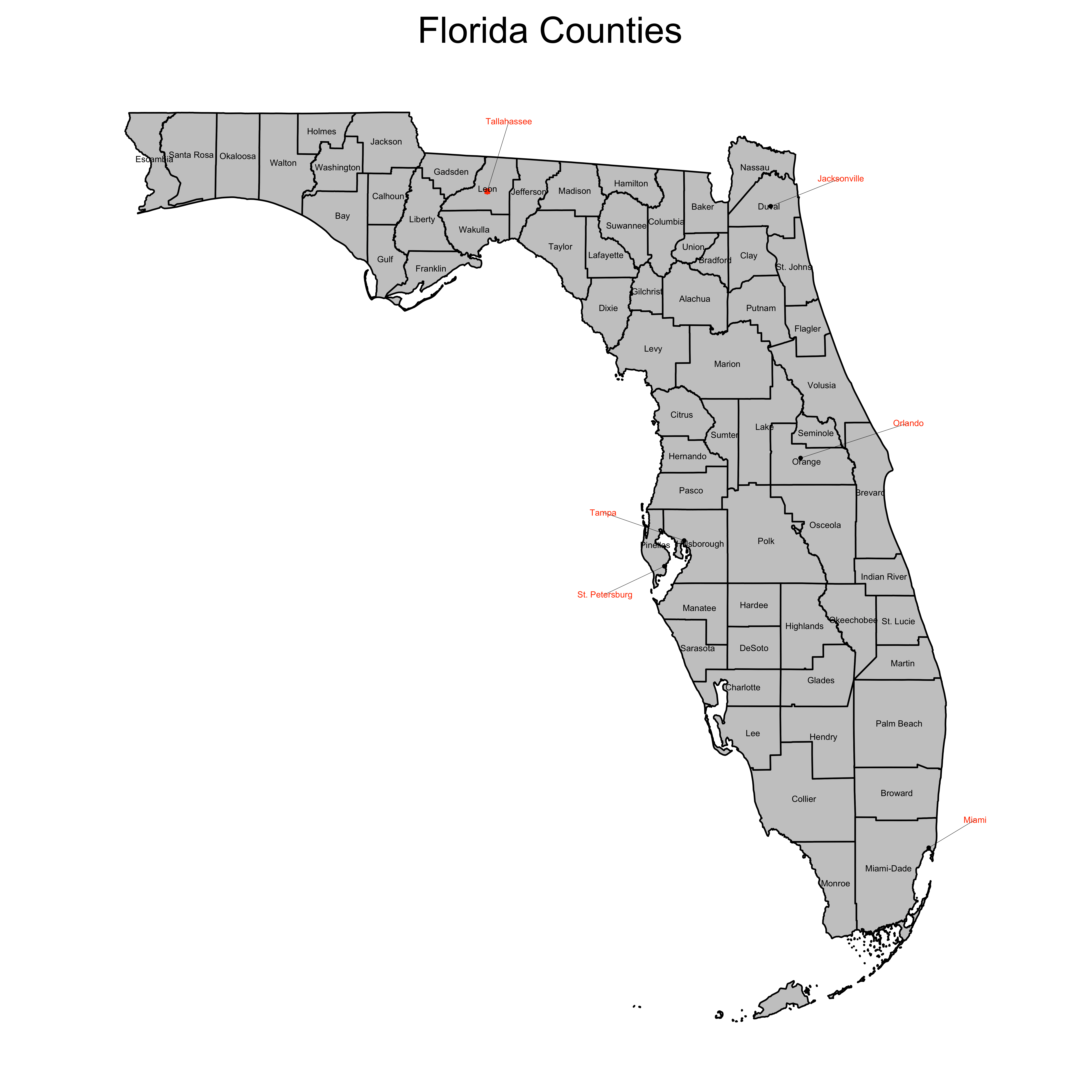

**Supplemental Figure 3**. Florida county map.
